# A methodology to generate longitudinally updated ACLF prognostication scores from electronic health record data

**DOI:** 10.1101/2020.11.23.20237081

**Authors:** Jin Ge, Nader Najafi, Wendi Zhao, Ma Somsouk, Margaret Fang, Jennifer C. Lai

**Author notes:** **Corresponding Author:** Jennifer C. Lai, MD, MBA, 513 Parnassus Avenue, UCSF Box 0538, San Francisco, CA 94143. **Abbreviations:** A ACLF, acute-on-chronic liver failure; CDR, clinical data repository; CI, confidence interval; CLIF-C-ACLF, European Association for the Study of Liver – Chronic Liver Failure Consortium – Acute-on-Chronic Liver Failure; CLIF-C-OF, European Association for the Study of Liver – Chronic Liver Failure Consortium – Organ Failures; CVVH, continuous venovenous hemofiltration; CVVHD, continuous venovenous hemodialysis; EF-CLIF, European Foundation for the Study of Chronic Liver Failure; EHR, electronic health record; ESLD, end-stage liver disease; FiO2, oxygen fraction; FrAILT, Multi-Center Functional Assessment in Liver Transplantation Study; GCS, Glasgow Coma Score; HE, hepatic encephalopathy; ICD, International Classification of Diseases; IQR, interquartile range; INR, international normalized ratio; LFI, Liver Frailty Index; MELD, Model for End-Stage Liver Disease; MELD-Na, Model for End-Stage Liver Disease-Sodium; NACSELD, North American Consortium for the Study of End-Stage Liver Disease; NACSELD-ACLF Score, North American Consortium for the Study of End-Stage Liver Disease – Acute-on-Chronic Liver Failure Score; NACSELD-OF, North American Consortium for the Study of End-Stage Liver Disease – Organ Failures; OMOP, Observational Medical Outcomes Partnership; P/F Ratio, partial pressure to fraction of oxygen ratio; RRT, renal replacement therapy; SpO2, oxygen saturation; SQL, Structured Query Language; WHC, West-Haven Criteria. **Financial Support:** This study was funded by 5T32DK060414-18 (National Institute of Diabetes and Digestive and Kidney Diseases, Ge) and R01AG059183/K23AG048337 (National Institute on Aging, Lai). The funding agency played no role in the analysis of the data or the preparation of this manuscript. **Author Contributions:** *Ge*: Study concept and design; analysis and interpretation of data; drafting of manuscript; critical revision of the manuscript for important intellectual content; statistical analysis *Najafi*: Acquisition of data; interpretation of data; critical revision of the manuscript for important intellectual content *Zhao*: Acquisition of data; interpretation of data *Somsouk*: Interpretation of data; critical revision of the manuscript for important intellectual content *Fang*: Acquisition of data; interpretation of data; critical revision of the manuscript for important intellectual content *Lai*: Study concept and design; analysis and interpretation of data; drafting of manuscript; critical revision of the manuscript for important intellectual content; obtained funding; study supervision. **Writing Assistance:** None.

## Abstract

**Background and Aims:** Queries of electronic health record (EHR) data repositories allow for automated data collection. These techniques have not been utilized in hepatology due to previous inability to capture hepatic encephalopathy (HE) grades, which are inputs for acute-on-chronic liver failure (ACLF) models. Here, we describe a methodology to utilizing EHR data to calculate rolling ACLF scores.

**Methods:** We examined 239 patient-admissions with end-stage liver disease 7/2014-6/2019. We mapped EHR flowsheet data to determine HE grades and calculated two longitudinally updated ACLF scores. We validated HE grades and ACLF diagnoses via chart review; and calculated sensitivity, specificity, and Cohen’s kappa.

**Results:** Of 239 patient-admissions analyzed, 37% women, 46% non-Hispanic White, median age 60 years, median MELD-Na at admission. Of the 239, 7% were diagnosed with NACSELD-ACLF at admission, 27% during the hospitalization, and 9% at discharge. Forty percent diagnosed with CLIF-C-ACLF at admission, 51% during the hospitalization, and 34% at discharge.

From chart review of 51 admissions, we found sensitivities and specificities for any HE (grades 1-4) were 92-97% and 76-95%, respectively; for severe HE (grades 3-4) were 100% and 78-98%, respectively. Cohen’s kappa between flowsheet and chart review HE grades ranged 0.55-0.72. Sensitivities and specificities for NACSELD-ACLF diagnoses were 75-100% and 96-100%, respectively; for CLIF-C-ACLF diagnoses were 91-100% and 96-100%, respectively. We generated approximately 28 unique ACLF scores per patient per admission-day.

**Conclusion:** In this study, we developed an informatics-based methodology for to calculate longitudinally updated ACLF scores. This opens new analytic potentials, such big data methods to develop electronic phenotypes for ACLF patients.

## Introduction

Electronic health records (EHRs) capture and generate vast amounts of granular clinical data through routine operations.(1) Structured Query Language (SQL) queries of associated clinical data repositories (CDRs) allow for automated generation of comprehensive laboratory, flowsheet, medical device, and medication administration reports for a cohort of patients.(2–4) Integration of these separate data reports have the potential to survey patients in a longitudinal fashion during inpatient admission and construct electronic phenotypes to define as subgroups of interest for further exploration.(5,6) Existing applications of SQL querying of data repositories in gastroenterology and hepatology, however, have been limited to searching International Classification of Diseases (ICD) 9/10 codes, identifying keywords in clinician documentation, and/or acquiring laboratory data.(7–9)

In hepatology research, specifically, the adoption of informatic methods described above has been hindered by inability to capture data to inform hepatic encephalopathy grades, which are often used as inputs into clinical prognostication models. Flowsheet reports, which contain structured and semi-structured entries reflecting interprofessional assessments of mentation, functional status, and physical exam findings, represent a rich source of relevant clinical information.(3,4) These semi-structured documentation of mentation and functional status have the potential to be mapped to describe hepatic encephalopathy, thereby enabling en-masse automated data acquisition for clinical research in hepatology.

This becomes especially relevant in the study of Acute-on-Chronic Liver Failure (ACLF), which is defined as the acute decompensation of end-stage liver disease (ESLD) with extrahepatic organ failures and high short-term mortality.(10–16) ACLF is a heterogenous and dynamic clinical syndrome with variable etiologies, triggers, and outcomes.(17) Reflecting the diversity of ACLF, several competing definitions and scoring systems currently exist, such as the North American Consortium for the Study of End-Stage Liver Disease (NACSELD diagnostic criteria and NACSELD-ACLF score)(10) and the European Association for the Study of Liver – Chronic Liver Failure Consortium (EF-CLIF diagnostic criteria and CLIF-C-ACLF score).(13) Existing ACLF prognostication scores, however, are generated in a cross-sectional manner at a specific point in time – thereby remain limited in clinical utility due to inconsistent abilities to predict recovery and identify transplant candidates.(18) Moreover, the lack of consensus on an unified prognostication model implies that standard methodologies for predictive modeling may be inadequate for this disease state. Longitudinally-updated ACLF scores, therefore, may be able to improve predictive ability and better inform ACLF outcomes research as previous studies have shown score changes and trajectories have greater prognostic value.(19–22)

In this study, we describe two-step methodology to generate longitudinally updated ACLF prognostication scores (NACSELD-ACLF and CLIF-C-ACLF scores):

1. Calculate West Haven Criteria (WHC) grades of hepatic encephalopathy by mapping mentation and functional status descriptors in flowsheet reports and validating these mapped WHC grades via chart review.
2. Integrate mapped WHC grades with relationally-linked reports of laboratory value, medical device data, and medication administration reports to generate longitudinally updated ACLF prognostication scores.

## Materials and Methods

We examined all inpatient admissions in a five-year period between July 1, 2014 through June 30, 2019 for the 1,918 patients enrolled in the Multi-Center Functional Assessment in Liver Transplantation (FrAILT) Study at a single academic medical center (University of California, San Francisco Medical Center) as of October 30, 2019. The FrAILT Study is a prospective, longitudinal study of adult patients with ESLD awaiting liver transplantation evaluated in the ambulatory care setting.(23) Hospital admissions for these patients were excluded if the admission took place after liver transplantation, was for a scheduled liver transplantation within 48 hours, with total length of study < 24 hours, or took place prior to their enrollment in the FrAILT Study. If a patient had multiple hospitalizations, we analyzed the hospitalization immediately after the most recent Liver Frailty Index (LFI) assessment to isolate one admission per patient (patient-admission). Of note, all patients who are listed for liver transplantation at our medical center are admitted to a dedicated multidisciplinary Liver Transplant Unit jointly attended by a hepatologist and a transplant surgeon for inpatient care. A flow diagram of the analyzed patient population from the FrAILT Study is shown in Figure 1.

**Figure 1.**
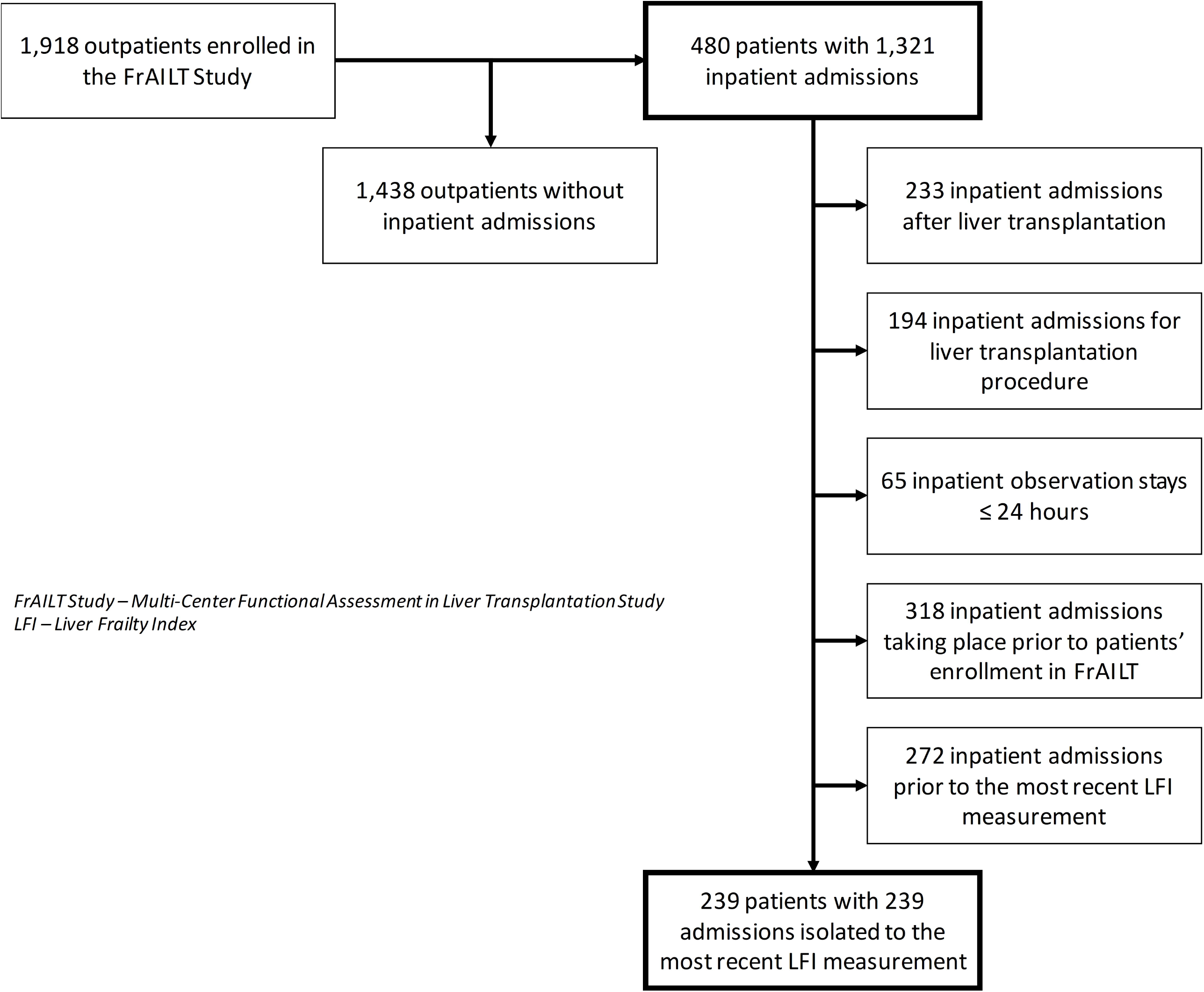
Isolation of the 239 Patient-Admissions Analyzed in this Study

Baseline demographic and clinical data were extracted from the date of the latest outpatient Liver Frailty Index assessment. Race/ethnicity was classified into the following categories: White, Black, Hispanic, Asian, Native American, or Other. Etiologies of liver disease were categorized as: chronic hepatitis C, alcohol-associated, autoimmune/cholestatic, chronic hepatitis B, and other etiologies. Patients were considered to comorbid diagnoses of hypertension, diabetes, or coronary artery disease if they were reported in the EHR. The Institutional Review Board at the University of California, San Francisco approved this study.

### Structured Query Language Data Collection

For this cohort of 1,918 patients and eligible admissions, we queried the EPIC (EpicCare, Epic Systems, Verona, WI) Clarity CDR hosted at the University of California, San Francisco Medical Center for the following reports containing data generated through routine care (Table 1). These reports were then linked relationally through two unique identifiers to each patient and admission: medical record number and contact serial number.

**Table 1.**
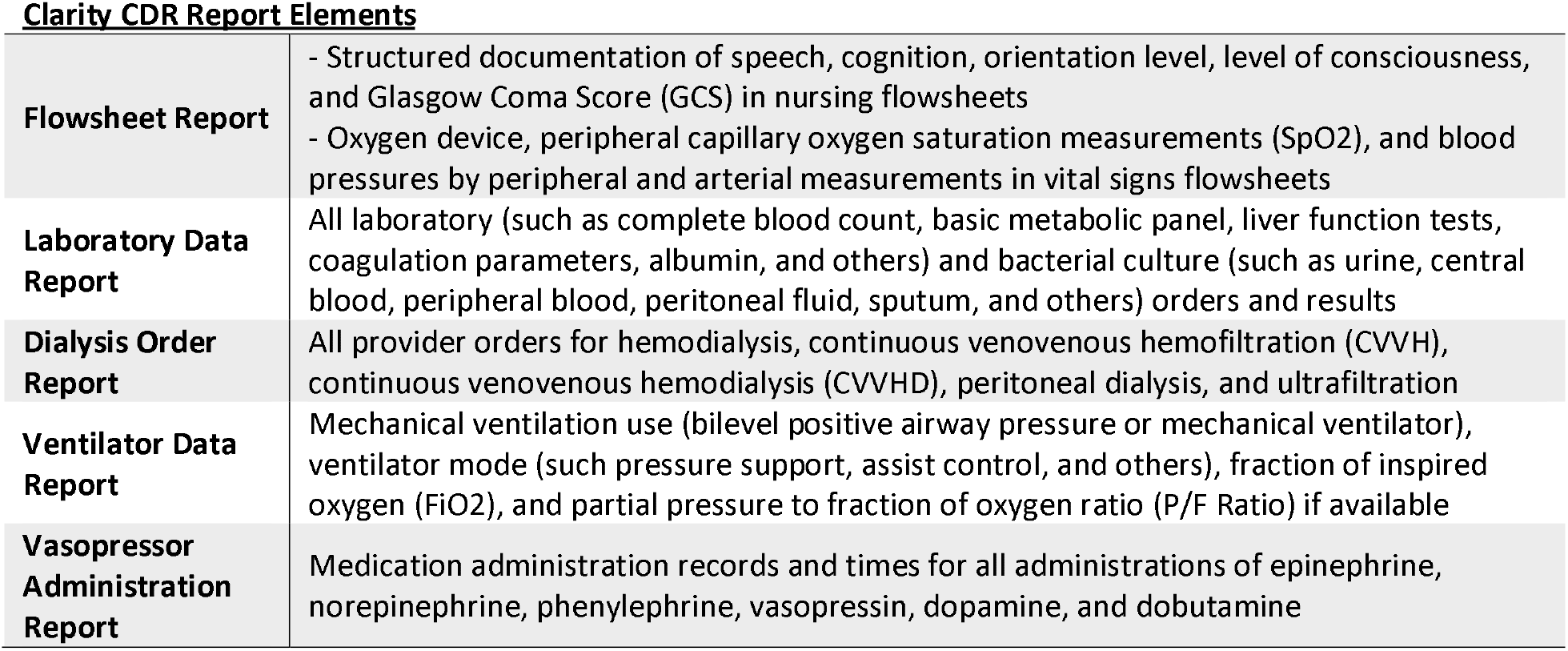
Data and Report Elements Generated from SQL Queries of Clarity CDR

### West Haven Criteria for Encephalopathy

Hepatic encephalopathy grades were mapped based on matching of standardized/structured entries for speech, cognition, orientation level, and level of consciousness in nursing flowsheets with descriptors used for grading HE per WHC with guidance from published criteria from Hepatic Encephalopathy Scoring Algorithm (Table 2).(24–27) GCS scores were also mapped with WHC grades with GCS 15 mapping to grade 0, GCS 12-14 mapping to grade 2, GCS 4-11 mapping to grade 3, and GCS 3 mapping to grade 4.(26) If there were multiple data entries (e.g. entries for GCS, level of consciousness, and orientation) recorded at a given time or if the entries mapped to discrepant WHC grades, the maximum mapped WHC grade was used by default to maximize detection sensitivity. Structured data entries that did not fall under the above criteria, such as “Other (comment),” and unstructured data entries were excluded from mapping and analysis.

**Table 2.** Mapping of Flowsheet Data on Four Domains of Mental Status and Glasgow Coma Score (GCS) to West Haven Criteria for Hepatic Encephalopathy*

| <u>West Haven Criteria</u><br><u>(WHC)(24,26,27)</u> | <u>Flowsheet Documentation of Mentation</u> |  |  |  | <u>Glasgow Coma</u><br><u>Score</u> |
| --- | --- | --- | --- | --- | --- |
|  | *Speech* | *Cognition* | *Orientation Level* | *Level of Consciousness* |  |
| **<u>Grade 0</u>** - "No encephalopathy at all, no history of HE." | • Clear<br>• Appropriate for developmental age<br>• Uses written communication<br>• Nods/gestures appropriately | • Appropriate judgment<br>• Appropriate safety awareness<br>• Appropriate attention/concentration<br>• Appropriate for developmental age<br>• Follows commands<br>• No short term memory loss | • Oriented x4<br>• Oriented to place<br>• Oriented to time<br>• Oriented to person<br>• Oriented to situation<br>• Appropriate for developmental age | • Alert<br>• Awake<br>• Responds to verbal | 15 |
| **<u>Grade 1</u>** - "Trivial lack of awareness, euphoria or anxiety, shortened attention span, impairment of addition or subtraction, altered sleep rhythm." | • Delayed responses | • Impulsive | • Disoriented to situation |  |  |
| **<u>Grade 2</u>** - "Lethargy or apathy, disorientation for time, obvious personality change, inappropriate behavior, dyspraxia, asterixis" | • Wordfinding difficulty<br>• Slurred | • Poor safety awareness<br>• Poor judgement<br>• Poor attention/concentration<br>• Short term memory loss | • Disoriented to person<br>• Disoriented to time | • Lethargic | 12-14 |
| **<u>Grade 3</u>** - "Somnolence to semi stupor, responsive to stimuli, confused, gross disorientation, bizarre behavior" | • Incomprehensible<br>• Expressive aphasia<br>• Receptive aphasia | • Unable to follow commands | • Disoriented to place<br>• Disoriented x4 | • Somnolent<br>• Responds to pain only<br>• Difficult to maintain arousal<br>• Confused | 4-11 |
| **<u>Grade 4</u>** - "Coma" | • Global aphasia |  |  | • Obtunded<br>• Unresponsive | 3 |

**\*This classification scheme was based on guidance from descriptors in the West Haven Criteria and criteria used in the Hepatic Encephalopathy Scoring Algorithm(24,26,27)**

To validate the mapped WHC grades, we conducted manual chart reviews of the relevant sections (subjective findings, physical examination, and assessment and plan) of history and physical notes, progress notes, and discharge summaries of a random subset (20%) of patient-admissions. WHC grades were assigned based on descriptors in the subjective and physical examination findings as matched to commonly accepted criteria described in practice guidelines(24), or as documented in the assessment and plan sections of the note. For example, physical examination findings of “lethargy” on chart review would be considered consistent with WHC grade 2 while findings of “arousable to voice only” or “grossly confused” would be consistent with WHC grade 3. If there were discrepancies between physical examination findings between different members of the provider team, then we utilized findings based on a hierarchical read based on the level of training. For example, the attending physician’s documented examination finding would be used over that of a resident physician etc. This physician review was conducted at three timepoints during each admission: at time of initial admission, during the hospitalization (defined as maximum value acquired during the admission), at the time of discharge.

### Determination of Oxygen Fraction (FiO2)

Whenever available, the recorded FiO2 in ventilator data or vital sign flowsheets reports were utilized. If such data were not available, then we estimated FiO2 based on nasal cannula and high flow nasal cannula flow rates assuming closed mouth breathing as previously validated in respiratory care literature (Supplemental Table 1).(28–30) These recorded and estimated FiO2 values were used to calculated SpO2/FiO2 and P/F ratios when appropriate.

### ACLF Definitions and Prognostication Score Calculation

For each patient-admission, we used to the above data and mapped WHC grades to diagnose ACLF and calculated prognostication scores based on those published by the North American Consortium for the Study of End-Stage Liver (NACSELD-ACLF score),(10) and European Foundation for the Study of Chronic Liver Failure (CLIF-C-ACLF score).(13) The NACSELD-ACLF score (range 0-1) predicts the probability of 30-day survival in hospitalized patients (Supplemental Table 2).(10) Model for End-Stage Liver Disease (MELD) and Model for End-Stage Liver Disease Sodium (MELDNa) scores were also calculated as previously described as inputs into the NACSELD-ACLF score.(31,32) Similarly, the CLIF-C-ACLF score is a composite score (range 0 to 100) with a score ≥ 70 predicting up to 100% mortality at 28 days (Supplemental Table 2).(13) We did not utilize the Asian Pacific Association for the Study of the Liver ACLF Research Consortium criteria for diagnosis due to our cohort being based in the United States and as ACLF etiologies differ significantly in patients based in the Asia-Pacific region.(18)

Using the mapped WHC grades from the methods above, we then generated automated diagnoses based on NACSELD and EF-CLIF criteria at the three timepoints specified for validation of WHC grades of HE: at time of initial admission, during the hospitalization (defined as maximum value acquired during the admission), at the time of discharge. To validate the automated ACLF diagnoses, we conducted physician chart reviews of the relevant sections (physical examination, laboratory findings, and assessment and plan) of history and physical notes, progress notes, and discharge summaries of the same random subset (20%) of patient-admissions validated in WHC grade validation. NACSELD and EF-CLIF ACLF diagnoses were confirmed based on descriptors in the physical examination findings, laboratory values, and diagnoses in the assessment and plan sections of the notes as matched to the relevant diagnostic criteria.(10,13)

### Statistical Analyses

Clinical characteristics and laboratory data for participants were summarized by medians and interquartile ranges (IQR) for continuous variables or numbers and percentages (%) for categorical variables. Comparisons among groups were performed using chi-square and Kruskal-Wallis tests where appropriate. We calculated sensitivities and specificities of the mapped WHC grades’ ability to detect any HE (WHC grades 1-4) and severe HE (WHC grades 3-4) uncovered by chart review. To rate interobserver agreement between mapped WHC grades and those acquired from chart review, we calculated Cohen’s kappa coefficients and generated 95% confidence intervals from bootstrapping with 1,000 replications.(33) Similarly, we also calculated sensitivities and specificities of the automated NACSELD-ACLF and EF-CLIF ACLF diagnoses versus manual chart diagnoses. Two-sided p-values <0.05 were considered statistically significant in all analyses. Analyses were performed using STATA statistical software, version 16.1 (StataCorp, College Station, TX, USA).

## Results

Of 1,918 patients in the FrAILT Study at the University of California, San Francisco; 480 patients (25%) had 1,321 admissions during the five-year study period. Of these 1,321 admissions, 233 occurred after liver transplantation, 194 were admissions for the liver transplantation surgical procedure, 65 lasting ≤ 24 hours, and 318 taking place prior to enrollment in the FrAILT Study. Of the 239 remaining patients meeting the inclusion criteria, we isolated one admission per patient for 239 patient-admissions (Figure 1).

### Baseline Characteristics

Baseline characteristics of the 239 patients are presented in Table 3: 37% were women, 46% were non-Hispanic White, and median age at admission was 60 years (IQR 53 to 65). The most common etiologies of cirrhosis were chronic hepatitis C (31%), alcohol-associated liver disease (24%), non-alcoholic fatty liver disease (21%), autoimmune/cholestatic diseases (10%), and chronic hepatitis B (5%). The median MELD upon admission was 21 (IQR 15 to 29) and the median MELD-Na upon admission was 25 (IQR 17 to 32).

**Table 3.**
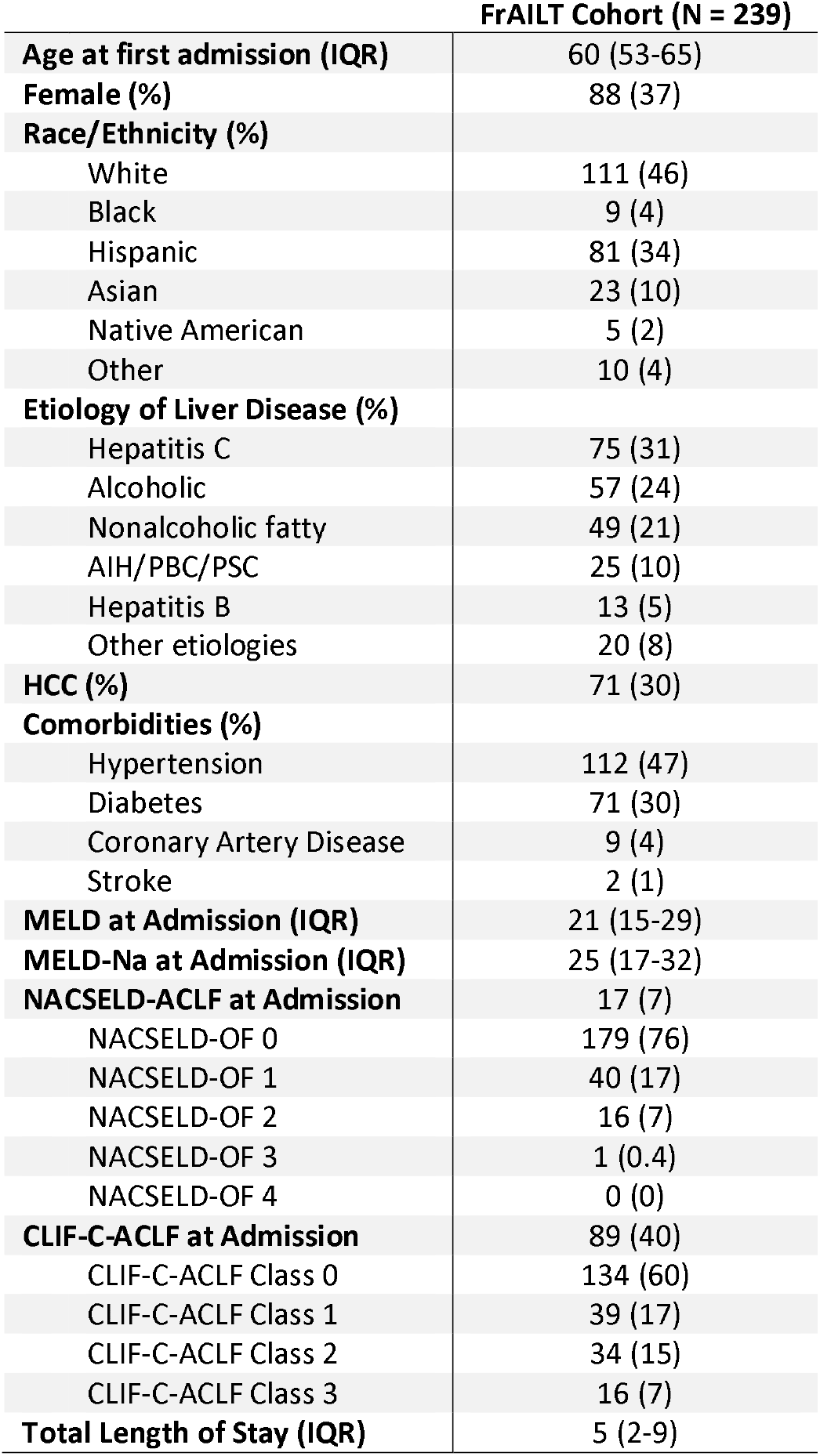
Baseline Clinical and Demographic Characteristics

### Validation of West Haven Criteria for Hepatic Encephalopathy

Given that validation of WHC grades using EHR has not previously been performed, we randomly selected 51 patient-admissions (21%) to undergo manual chart review to validate the WHC grades mapped from flowsheet data. Sensitivities and specificities (along with 95% confidence intervals) for presence of any HE and severe HE by comparing mapped WHC grades versus chart review are presented in Table 4. The sensitivities for the presence of any HE ranged from 92-97% while those for severe HE was 100% at the three timepoints queried (the initial time of admission, during hospitalization, and the time of discharge). Specificities for the presence of any HE ranged from 76-95% while those for severe HE ranged from 78-98% at the three timepoints queried. Cohen’s kappa coefficient for agreement between different WHC grades were 0.55 (95%CI 0.33-0.74) at time of admission, 0.64 (95%CI 0.49-0.79) at the time of maximum value in the middle of the admission, and 0.72 (95%CI 0.51-0.90) at time of discharge.

**Table 4.** Sensitivity and Specificity of Mapped WHC versus Chart Review

|  | True Positive | False Negative | False Positive | True Negative | Sensitivity | 95% CI | Specificity | 95% CI |
| --- | --- | --- | --- | --- | --- | --- | --- | --- |
| Any HE (WHC 1-4) at Admission | 12 | 1 | 9 | 29 | 0.92 | 0.64-1.00 | 0.76 | 0.60-0.89 |
| Any HE (WHC 1-4) during Hospitalization | 31 | 1 | 3 | 16 | 0.97 | 0.84-1.00 | 0.84 | 0.60-0.97 |
| Any HE (WHC 1-4) at Discharge | 11 | 1 | 2 | 37 | 0.92 | 0.61-1.00 | 0.95 | 0.83-0.99 |
| Severe HE (WHC 3-4) at Admission | 4 | 0 | 6 | 41 | 1.00 | 0.40-1.00 | 0.87 | 0.74-0.95 |
| Severe HE (WHC 3-4) during Hospitalization | 15 | 0 | 8 | 28 | 1.00 | 0.78-1.00 | 0.78 | 0.61-0.90 |
| Severe HE (WHC 3-4) at Discharge | 5 | 0 | 1 | 45 | 1.00 | 0.48-1.00 | 0.98 | 0.89-1.00 |

### ACLF Diagnoses

For the NACSELD-ACLF diagnostic criteria for ACLF, 17 patients (7%) were diagnosed at the time of initial admission with 16 having two organ failures and one having three organ failures. The number of patients diagnosed with ACLF by the NACSELD criteria increased to 64 (27%) during the hospitalization with 20 having two, 18 having three, and 26 having four organ failures. At the time of discharge, 21 (9%) still had ACLF diagnoses with 7 having two, 6 having three, and 8 having four organ failures. In comparison to manual chart review of the same 51 patient-admissions selected for WHC validation, we found that the sensitivities and specificities for ACLF diagnoses under the NACSELD criteria to be 75% and 96%, respectively, at the time of initial admission. The sensitivity and specificities increased to 100% and 97%, respectively, during the admission; and then to 100% and 100%, respectively, at the time of discharge (Table 5).

**Table 5.** Sensitivity and Specificity of Calculated NACSELD-ACLF and CLIF-C-ACLF Diagnoses versus Chart Review

|  | True Positive | False Negative | False Positive | True Negative | Sensitivity | 95% CI | Specificity | 95% CI |
| --- | --- | --- | --- | --- | --- | --- | --- | --- |
| Dx of NACSELD-ACLF at Admission | 3 | 1 | 2 | 45 | 0.75 | 0.19-0.99 | 0.96 | 0.86-1.00 |
| Dx of NACSELD-ACLF during Hospitalization | 15 | 0 | 1 | 36 | 1.00 | 0.78-1.00 | 0.97 | 0.86-1.00 |
| Dx of NACSELD-ACLF at Discharge | 4 | 0 | 0 | 47 | 1.00 | 0.40-1.00 | 1.00 | 0.93-1.00 |
| Dx of CLIF-C-ACLF at Admission | 21 | 2 | 1 | 27 | 0.91 | 0.72-0.99 | 0.96 | 0.82-1.00 |
| Dx of CLIF-C-ACLF during Hospitalization | 27 | 0 | 0 | 24 | 1.00 | 0.87-1.00 | 1.00 | 0.86-1.00 |
| Dx of CLIF-C-ACLF at Discharge | 19 | 0 | 0 | 32 | 1.00 | 0.82-1.00 | 1.00 | 0.89-1.00 |

With respect to the EF-CLIF ACLF diagnostic criteria, 89 patients (40%) wee diagnosed at the time of initial admission with 39 meeting grade one, 34 grade two, and 16 grade three criteria. The number of patients diagnosed with ACLF by the EF-CLIF criteria increased to 114 (51%) during the hospitalization with 28 meeting grade one, 34 grade two, and 52 grade three criteria. At the time of discharge, 76 (34%) still had ACLF diagnoses with 35 meeting grade one, 21 grade two, and 19 grade three criteria. Similarly, when we compared to manual chart review of the same 51 patient-admissions selected for WHC validation, we found that the sensitivities and specificities for ACLF diagnoses under the EF-CLIF criteria to be 91% and 96%, respectively, at the time of initial admission. These figures increased to 100% and 100%, respectively, during the admission and at the time of discharge (Table 5).

### Longitudinal ACLF Prognostication Scores

A total of 44,639 unique data points from the 239 patient-admissions were available for ACLF prognostication score generation. This represented a median 454 data points per admission (IQR 194 to 704) and median 28 data points per admission-day (IQR 21 to 35). Using the data points generated from the relationally-linked databases, we were able to calculate approximately hourly updated NACSELD-ACLF and CLIF-C-ACLF scores. A representative example of one patient’s hospitalization course, and the corresponding NACSELD-ACLF and CLIF-C-ACLF scores, is shown in Figure 2.

**Figure 2.**
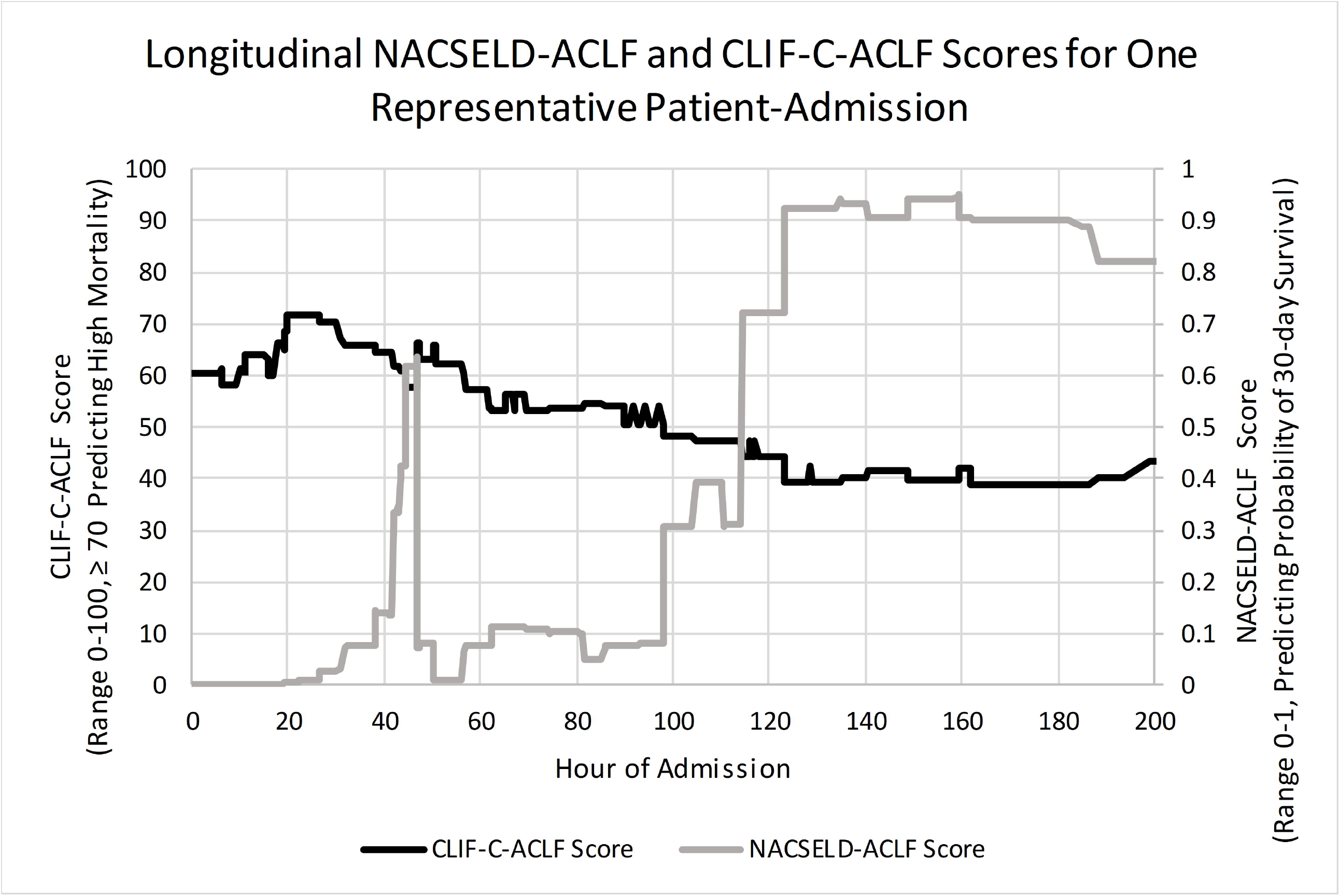
Representative Sample of Calculated Longitudinal NACSELD-ACLF and CLIF-C-ACLF Scores

## Discussion

In this study, we validated an informatics-based method for capturing routine clinical care data from the electronic health record to calculate longitudinally updated ACLF prognostication scores in patients with ESLD hospitalized for ACLF. We extracted and mapped flowsheet data to WHC grades for HE and integrated these mapped values to more traditional data (laboratory values, device data, and medication administration records) to diagnose ACLF and calculate longitudinally-updated prognostic scores under two definitions (NACSELD-ACLF and CLIF-C-ACLF scores, Figure 2). Our method demonstrated high sensitivity in detecting any or severe HE throughout the hospitalization, and moderate sensitivity for diagnosing ACLF at admission that rapidly improved during the hospitalization.

While the techniques of using SQL queries of CDRs have been previously demonstrated,(1) existing applications have been limited.(7–9) Our method is novel in that it integrates multiple sources of EHR data: laboratory data, medication administration records, provider orders, ventilator device data, and flowsheet data. We made extensive use of flowsheets, which contain interprofessional (particularly nursing) assessments of mentation, functional status, and examination that contained structured data entries that mapped to WHC grades based on previously validated instruments. Flowsheet data comprise 1/3 of all recorded data in CDRs and have been historically underutilized.(3,4) Indeed, the linchpins in our novel methodology were mapping of flowsheet entries to appropriate WHC grades and of supplemental oxygenation flow rates to estimated FiO2.

Validation of WHC grades generated from flowsheet data proved to have high sensitivity (92-95% for any HE and 100% for severe HE) versus clinicians’ documentation. Of note, HE grading has historically been difficult due to subjective assessments with poor to moderate interrater reliability.(24,34–37) This appears to be the rationale behind the use of overt hepatic encephalopathy, which have has interrater reliability, for diagnosing brain failure in both NACSELD and EF-CLIF definitions.(34) Our calculations of Cohen’s Kappa coefficient indicated moderate agreement between WHC grades mapped from flowsheet data and those rated by clinicians on retrospective chart review. The Kappa coefficients from our study (0.55-0.72) are within range of those previously reported for other methodologies for differentiating WHC grades.(26,38,39) While the sensitivities and specificities of our methodology for ACLF diagnosis (based on NACSELD and EF-CLIF criteria) were imperfect at the time of initial admission, they are rapidly improved to 100% as more information was generated and gathered throughout the admission.

We acknowledge the following limitations to our study. The first is that our methodology was developed at a single-center and on a specific implementation of the EPIC EHR system. Given that our patient population are largely cared for on a dedicated Liver Transplant Unit, our staff members may be more attuned to documenting mentation accurately versus other nursing units. Specific data elements recorded in flowsheets will likely differ between institutions, but interprofessional documentation reflected flowsheet elements are generally standardized by the Joint Commission and professional associations, such as the American Nursing Association.(4,40) This is a major advantage of our methodology in that it utilizes existing interdisciplinary training and charting infrastructure to discern different gradations of hepatic encephalopathy. In addition, while the exact execution for this methodology will differ at another institution, the general strategy of mapping and extracting flowsheet data remains the same. In our experience, while SQL code written for our institution’s CDR does not often work “out-of-the-box” against that of another institution – the differences are generally correctable and reconcilable. Reconciliation of flowsheet data elements in a multicenter setting has been previously demonstrated with regards to the creation of a pain information model.(41) In addition, the movement towards “standard” data models, such as the Observational Medical Outcomes Partnership (OMOP), will likely greatly increase portability in the future.(42)

Second, the patient population evaluated in this study was highly selected – we only considered patients with ESLD who were enrolled in the FrAILT Study (evaluated for transplantation). This high degree of selection, however, was by design to validate our methodology in a controlled cohort and raise possibilities for implementations in larger cohorts, such as all patients with ICD-9/10 discharge diagnoses of cirrhosis. Lastly, given that the University of California, San Francisco Medical Center is a tertiary referral center, many of the admissions evaluated in our study were transferred to our medical center. Patient-admissions in our sample, thus, may not reflect the initial clinical course. Moreover, we suspect that initial delays in documentation or clinician order placement (such as entering orders for dialysis) for transfer admissions contributed to the relatively poor sensitivity of our method for diagnosis of NACSELD ACLF upon admission compared to chart review. As expected, the accuracy and precision of our methodology increased through the length of stay as more data is integrated.

Despite these limitations, this study serves as a proof of concept for a clinical informatics-based methodology to generate longitudinally updated ACLF prognostication scores, which can better reflect the dynamic clinical course of these patients. Pilot demonstration of the validity of this methodology to extract accurate data in this population opens new analytic potentials, such as the application of big data methods, that leverage the rich data from EHR platforms and CDR configurations to enhance investigation of predictors of outcomes in this dynamic population.

## Data Availability

The data that support the findings of this study are available on request from the corresponding author. The data are not publicly available due to privacy or ethical restrictions.

---

Supplemental Table 1 – Conversion of Nasal Cannula and High-Flow Nasal Cannula Flow Rates to Estimated FiO2

Supplemental Table 2 – Diagnoses of ACLF based on NACSELD and EF-CLIF Criteria

