## Supplemental Data 1 for "A methodology to generate longitudinally updated ACLF prognostication scores from electronic health record data"

**Supplemental Table 1 – Conversion of Nasal Cannula and High-Flow Nasal Cannula Flow Rates to Estimated FiO2**

| Flow (L/min) | Estimated FiO2* |
| --- | --- |
| 1 | 0.24 |
| 2 | 0.30 |
| 3 | 0.35 |
| 4 | 0.40 |
| 5 | 0.45 |
| 6 | 0.48 |
| 7 | 0.51 |
| 8 | 0.50 |
| 9 | 0.56 |
| 10 | 0.59 |
| 15 | 0.80 |
| 20 | 0.90 |
| >25 | 0.95 |
| *Based on nasal cannula and high flow nasal cannula flow rates assuming closed mouth breathing as previously validated.(1–3) | |
