## Supplemental Data 2 for "A methodology to generate longitudinally updated ACLF prognostication scores from electronic health record data"

**Supplemental Table 2 – Diagnoses of ACLF based on NACSELD and EF-CLIF Criteria**

| NACSELD Criteria for Diagnosis of ACLF (10) | | EF-CLIF Criteria for Diagnosis of ACLF (13) | |
| --- | --- | --- | --- |
| NACSELD Organ Failure Definitions | | **EF-CLIF Organ Failure Definitions** | |
| *Brain Failure* | West-Haven Criteria grades 3-4 derived from flowsheet data | ***Brain Failure*** | West-Haven Criteria grades 3-4 derived from flowsheet data |
| *Renal Failure* | Need for renal replacement therapy (RRT) derived from dialysis order reports | ***Renal Failure*** | Serum creatinine ≥ 2mg/dl by laboratory data report or need for RRT from dialysis order reports |
| *Respiratory Failure* | Need for bilevel positive airway pressure or mechanical ventilation based on flowsheet oxygenation reports and ventilator data reports | ***Respiratory Failure*** | SpO2/FiO2 ratio of ≤ 214 based on peripheral capillary oxygen saturation from vital signs flowsheet reports |
| *Shock* | Need for vasopressors based on medication administration reports and a mean arterial pressure of < 60mmHg | ***Circulatory Failure*** | Need for vasopressors based on medication administration reports and a mean arterial pressure of < 60mmHg |
|  |  | ***Liver Failure*** | Total bilirubin ≥ 12 mg/dl by laboratory data report |
|  |  | ***Coagulation Failure*** | International normalized ratio (INR) ≥ 2.5 by laboratory data report |
| NACSELD ACLF Diagnosis | Based on two or more organ failures above (NACSELD-OFs) | **EF-CLIF ACLF Diagnosis** | *Grade 1* - Presence of single kidney failure or any other organ failure when in combination with either renal insufficiency (serum creatinine ≥ 1.5 mg/dl) or WHC grades 1-2 *Grades 2 or 3* - Defined by the presence of two or at least three organ failures, respectively |
| NACSELD-ACLF Score | Θ/(1 + Θ), where Θ = exp(6.89 – 1.74*NACSELD-ACLF Diagnosis – 0.048*Age – 0.056*WBC + 0.31*Albumin – 0.085*MELD) | **CLIC-C-ACLF Score** | 10*(0.33*CLIF-C-OFs + 0.04*Age + 0.63*ln(WBC) – 2) |
